# Evaluating the effectiveness of social distancing interventions against COVID-19

**DOI:** 10.1101/2020.03.27.20044891

**Authors:** Laura Matrajt, Tiffany Leung

## Abstract

SARS-CoV-2 has infected over 140,000 people as of March 14, 2020. We use a mathematical model to investigate the effectiveness of social distancing interventions lasting six weeks in a middle-sized city in the US. We explore four social distancing strategies by reducing the contacts of adults over 60 years old, adults over 60 years old and children, all adults (25, 75 or 95% compliance), and everyone in the population. Our results suggest that social distancing interventions can avert cases by 20% and hospitalizations and deaths by 90% even with modest compliance within adults as long as the intervention is kept in place, but the epidemic is set to rebound once the intervention is lifted. Our models suggest that social distancing interventions will buy crucial time but need to occur in conjunction with testing and contact tracing of all suspected cases to mitigate transmission of SARS-CoV-2.

## 1 Introduction

Since the emergence of the novel coronavirus SARS-CoV-2 in Wuhan, China in December 2019 [1], the World Health Organization has declared the coronavirus disease 2019 (COVID-19) a pandemic [2]. As of March 14, 2020, COVID-19 has spread to over 114 countries, with 142,539 confirmed cases and 5,393 deaths related to COVID-19 globally [3].

On January 21, 2020, the first case of COVID-19 in the United States (US) was identified from a traveler who had recently returned to Snohomish County, Washington from Wuhan, China [4, 5]. As of the afternoon of March 14, there are 642 confirmed cases and 40 deaths associated with COVID-19 in Washington state [6]. In response to the rapid spread of COVID-19, on March 12, 2020 (approximately seven weeks after the first confirmed case in Washington state), the governor of Washington announced a state-wide prohibition of large gatherings (of 250 people or more) [7] and school closures lasting six weeks (from March 17 to April 24, 2020) in three counties [8]. Three days after, on March 15, the governor of Washington further issued a temporary shutdown of restaurants, bars, and entertainment and recreational facilities [9]. Similar interventions have now been enacted in several states in the US [10] and in countries in Europe [11, 12].

In this paper, we use an epidemic mathematical model to quantify the effectiveness of social distancing interventions in a medium-sized city in the US or Europe by taking Seattle, Washington, USA as an example. We provide estimates of the proportion of cases, hospitalizations, and deaths averted in the short term and identify key challenges in evaluating the effectiveness of these interventions.

## 2 Methods

We developed an age-structured susceptible-exposed-infectious-removed (SEIR) model to describe the transmission of COVID-19. We divided the population into ten age groups: 0–5, 6–9, 10–19, 20–29, 30–39, 40–49, 50–59, 60–69, 70–79, and *>* 80 years old (yo). We calibrated the model to the Seattle, WA metropolitan area and the age distribution of the population using census data [13]. Namely, for each age group, the population is divided into compartments: susceptible *S*, exposed *E* (those who have been infected but are not yet infectious), infectious *I* and removed *R* (Figure 1). Table 1 gives the parameters and ranges used in the model. Based on current estimates [14] that put less than one percent of the infections as asymptomatic, we considered only symptomatic infections. We used the contact matrix for six age groups computed by Wallinga et al. [15] and extend it to ten age groups. A full description of the model can be found in the Appendix.

**Table 1:** Description of parameters used in the SEIR model.

| Parameter | Meaning | Value | Range | Reference |
| --- | --- | --- | --- | --- |
| $1/\sigma$ | mean latent period | 5.16 d | 4.5–5.8 | [17] |
| $1/\gamma$ | mean infectious period | 5.02 d | 3–9 | [18] |
| $\beta$ | transmission coefficient | calculated | — | — |
| $\mathcal{C}$ | contact matrix | — | — | [15] |
| $N$ | Total population | 3.5 million | — | [13] |

**Figure 1:**
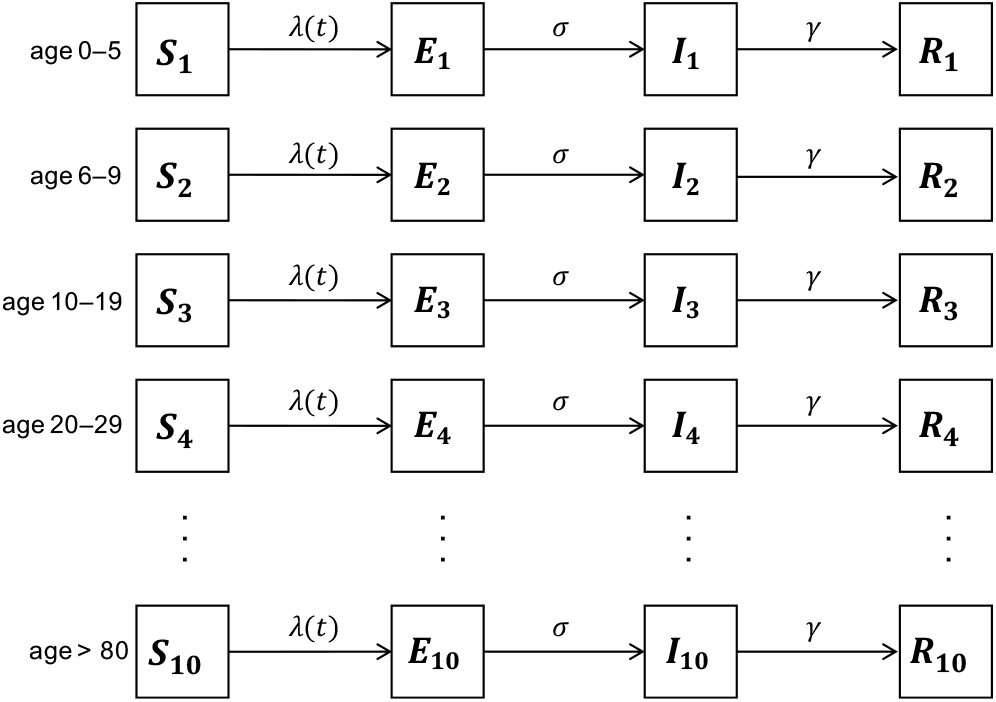
Model diagram illustrating the population divided into ten age groups, and for each age group (represented by subscripts) into compartments susceptible *S*, exposed *E*, infectious *I*, and removed *R*. Susceptible individuals become exposed at the force of infection *λ*(*t*), progress to become infectious at rate *σ*, and are removed from infecting others at rate *γ*.

Using genomic epidemiology of the first two cases identified in the state of Washington, Bedford found that by the time that the second case was identified in Washington state, SARS-CoV-2 had been circulating locally for six weeks [16]. Hence, we considered January 21, 2020 (the day the first case was identified) to be the first day of our simulation.

We assumed that 20% of the cases would be observed, as 80% of the cases are reported to be mild and would probably be missed [19]. Case fatality rates were obtained from [19] and hospitalization rates by age group were obtained from [20]. We modeled social distancing by reducing the contact rates in a given group for six weeks, corresponding to the policy in Washington state in mid-March [8, 21]. We divided the population into three major groups for the purposes of social distancing interventions: children (0–19 yo), adults under 60 yo (20–59 yo), and adults over 60 yo.

We investigate the effectiveness of four scenarios of social distancing: 1) Adults over 60 yo only, where contacts of adults over 60 yo are reduced by 95%; 2) Adults over 60 and children, where contacts of adults over 60 yo are reduced by 95%, and contacts of children are reduced by 85%; 3) Adults only, where contacts of adults over 60 yo are reduced by 95%, and contacts of adults under 60 yo by 25, 75, or 95%; and 4) All, where contacts are reduced for every group (for adults over 60 yo by 95%, for children by 85%, and for adults under 60 yo by 25, 75, or 95%).

To quantify the uncertainty around our results, we performed 1,000 simulations where we varied three parameters: the basic reproduction number *R*_0_, the latent period and the duration of infectiousness while keeping the other parameters fixed. Based on current estimates [1, 22, 23, 24], we sampled values of *R*_0_ from a truncated normal distribution with mean 2.26 ranging from 1.6 to 3. We assumed that the latent period was similar to the incubation period, and assumed a gamma distribution with mean 5.1 days and standard deviation 0.7 [17]. We sampled the duration of infectiousness from a truncated normal distribution with mean 5 days and standard deviation 0.7 ranging from 3 to 9 [18, 25]. For each *R*_0_, infectious period and latent period considered, we used the next-generation matrix approach [26] to calculate the transmission coefficient *β*. For each statistic presented in the results, we computed the error bars by removing the top and bottom 2.5% of the simulations.

## 3 Results

Given the current uncertainty surrounding the duration of infectiousness (with estimates ranging from 5.5 days [18] to as long as 20 days [27]), we analyzed its influence on the effectiveness of the social distancing interventions. We considered an epidemic with infectious periods of 5, 6, 7, or 8 days (corresponding to the most plausible values [18, 25], with all other parameters fixed. When the infectious period is shorter (5 days) the epidemic peaks at 85 days after the introduction of the first case. As the infectious period becomes longer, the epidemic takes much longer to take off (Figure 2). This is because we are keeping a fixed *R*_0_, so a longer infectious period implies a smaller infectious rate. Indeed, with the longest infectious period, the epidemic peaks 110 days after the first case was introduced. Hence, interventions occurring early on delay the epidemic, but do not substantially change the pool of susceptible individuals, allowing similar-sized epidemics to occur later (Figure 2).

**Figure 2:**
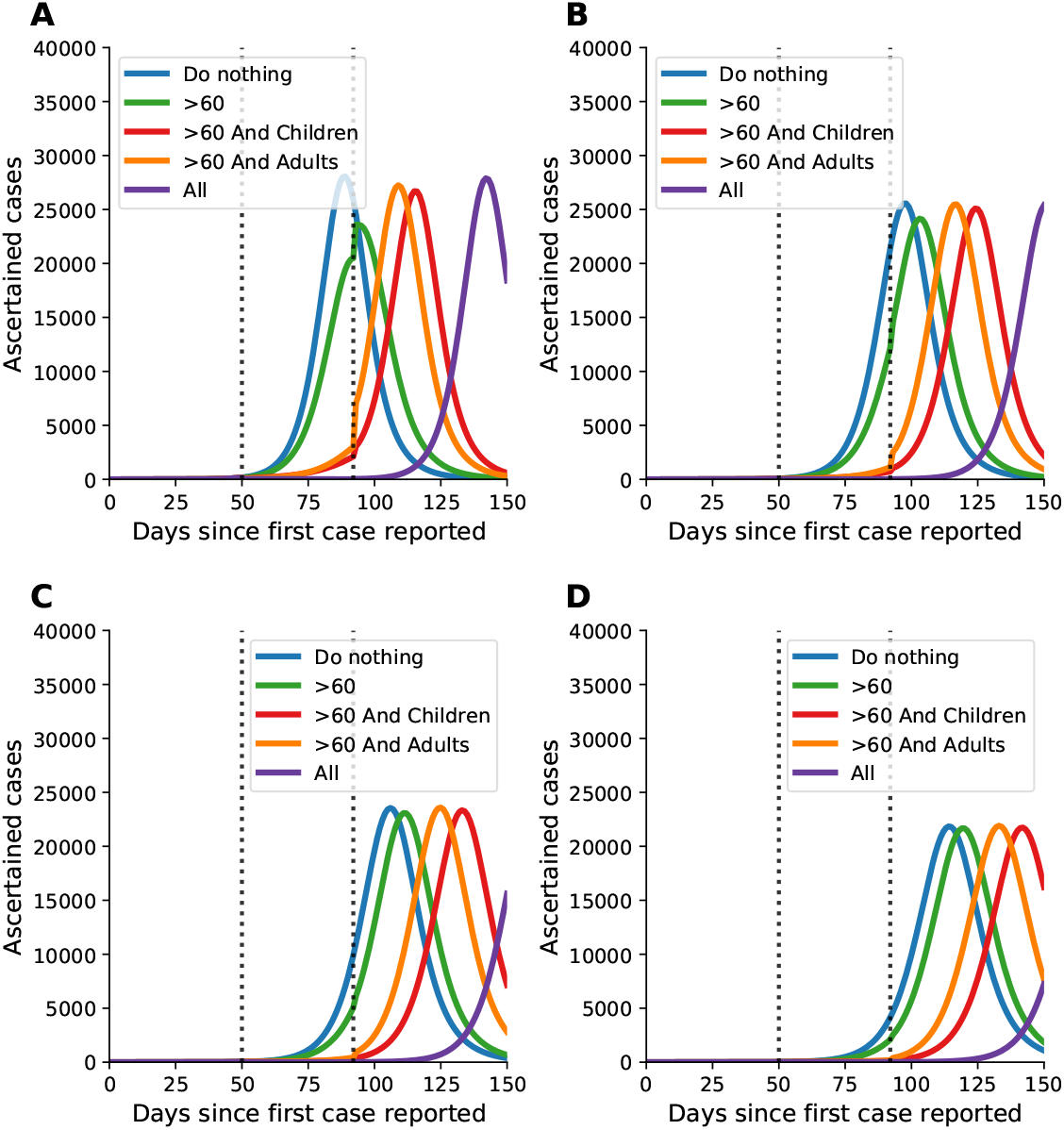
Number of ascertained cases over 150 days for varying infectious periods of (A) 5 days, (B) 6 days, (C) 7 days, and (D) 8 days (parameter values used: *R*_0_ = 2.26, *γ* = 1*/*5.02, *σ* = 1*/*5.16, and a 75% reduction of contacts in adults). The dotted lines represent the beginning and end of the social distancing intervention.

We then considered the delay of the epidemic under the four social distancing interventions and different infectious periods (Figure 2). As expected, the social distancing strategy applied to all delays the epidemic the most, by over 40 days compared to baseline (do nothing). Targeting adults over 60 and children delays the epidemic by 23 days irrespective of infectious period, and targeting adults and adults over 60 gives a delay of 16 to 19 days (for infectious period of 8 and 5 days, respectively). Social distancing of adults over 60 only delays the epidemic no more than 7 days irrespective of infectious period (Table S1). Overall, social distancing interventions under a shorter infectious period result in a longer delay of epidemic peak. The infectious period did not have significant effect to the peak height of the epidemic relative to baseline.

We examine the effectiveness of the three social distancing interventions in adults. Figure 3, panels A, B, C show the number of ascertained cases when the contact of adults are reduced by 25%, 75%, and 95%, respectively. Further, we consider two dates to start the social distancing interventions, at 50 (Figure 3, panels A–C) and 80 days (Figure 3, panels D–F). The effect of these interventions is dramatically different if started early relative to the epidemic curve (before the exponential phase of the epidemic) or later.

**Figure 3:**
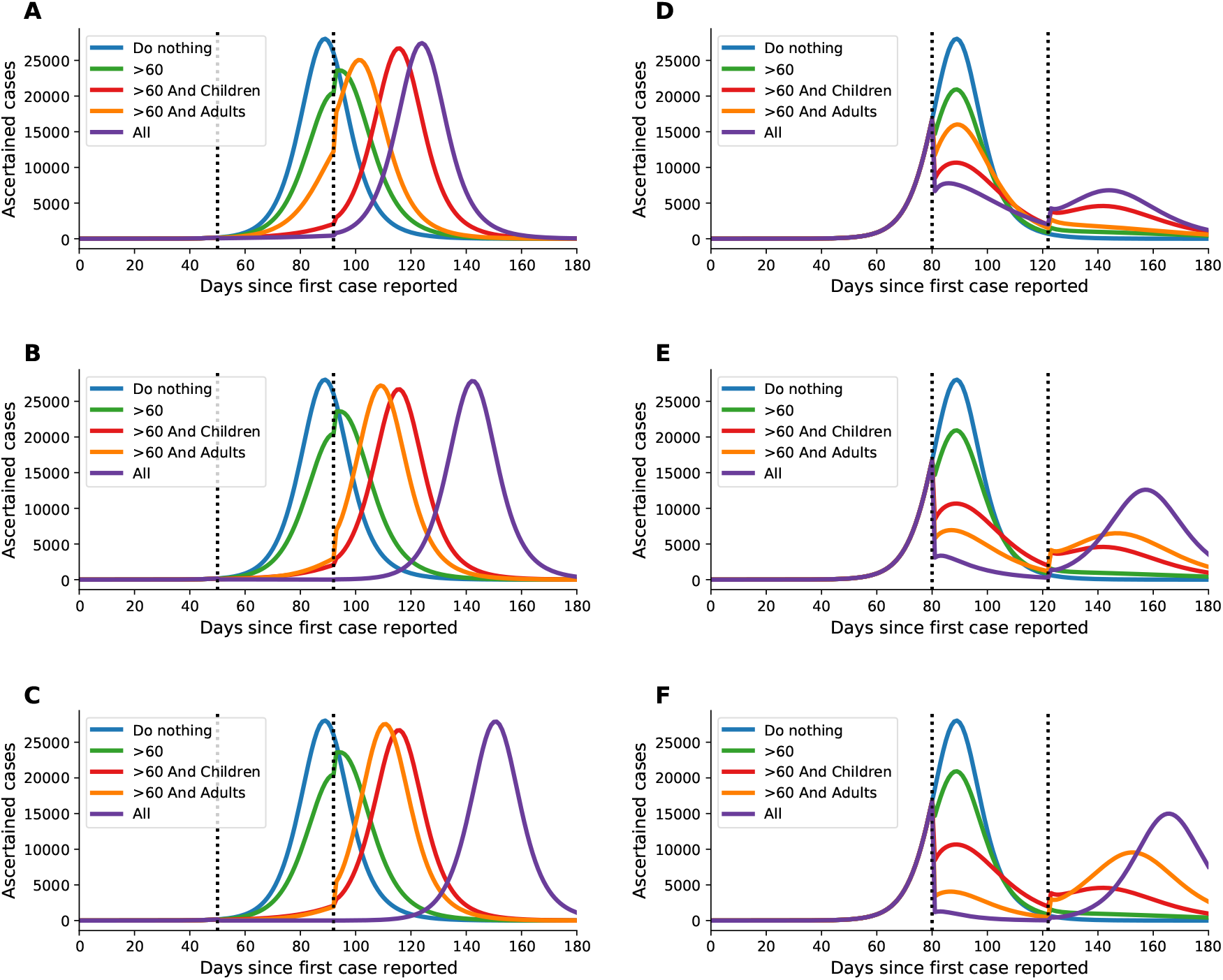
Number of ascertained cases over time with adults reducing their contact by (A) 25%, (B) 75%, and (C) 95% (parameter values used: *R*_0_ = 2.26, *γ* = 1*/*5.02, *σ* = 1*/*5.16). The dotted lines represent the beginning and end of the social distancing intervention. For panels A–C, intervention starts at day 50; for panels D–F, intervention starts at day 80. Social distancing interventions last 6 weeks before contact rates return to normal.

When interventions start on day 50, there is a delay of the epidemic regardless of the level of compliance in the adult population, with little change in the magnitude of the epidemic peak (a thorough analysis of this scenario is presented below). In comparison, if the intervention begins later (in the exponential phase of the epidemic), all interventions result in a flatter epidemic curve. The strategy of reducing the contacts of only the older adults results in a moderate reduction in the epidemic peak (7,000 fewer cases at the epidemic peak compared to baseline). It is the only intervention that does not exhibit a rebound once the intervention is lifted (Figure 3, panels D–F).

The strategy that targets adults over 60 and children results in 16,500 fewer cases (57% less than baseline) at epidemic peak (Figure 3, panels D–F) compared to baseline. In comparison, the adults-only strategy yields 12,000 fewer ascertained cases (41% less than baseline) at the epidemic peak for 25% compliance in younger adults (adults under 60 yo, Figure 3, panel D). At a higher compliance of 75% and 95% in adults, the epidemic peak is reduced even more, with 21,000 and 24,000 fewer cases ascertained at the peak (72 and 82% less) respectively (Figure 3, panel E, F), and the epidemic curve grows at a slower rate. Of the four different interventions, the all-groups strategy, which reduces the contacts in all age groups, decreases the epidemic peak the most and results in the slowest growth rate, as expected. Even with low compliance (25%) in the younger-adult groups, the all-groups strategy yields 20,000 fewer cases ascertained (68% less) at the epidemic peak (Figure 3, panel D). With higher compliance (95%) in adults, the all-groups strategy results in even fewer cases ascertained at the peak (27,000 fewer cases or 92% less; Figure 3, panel F). However, our results suggest that all these interventions can result in new epidemic curves once the interventions are lifted.

For this part of the analysis, we consider the effects of social distancing interventions over the first 100 days of the epidemic and assume that the social distancing interventions started on day 50. This corresponds to the approximate date where social distancing interventions started in Washington state. To investigate the sensitivity of the model to the parameters chosen, we ran 1,000 simulations (as described in the Methods). We obtained curves that varied widely over both the number of cases and the duration and timing of the epidemic (figures S1, S2, S3). We show here results of simulations done with the mean values (*R*_0_ = 2.26, an infectious period lasting 5 days and a latent period of 5.1 days). We report cases as well as the proportion of cases, hospitalizations and deaths averted during the first 100 days. While reducing the contacts of adults over 60 only averts only a moderate number of cases (21%, Figure 3; and 57% for the adults over 60, Figure S4)), the number of hospitalizations and deaths are reduced by 36% and 47% respectively for the whole population (Figure 4), and by 56% for the adults over 60 yo (Figure S5). Adding a social distancing intervention in children drastically slows down the epidemic curve (Figure 3). Further, hospitalizations and deaths are reduced by 75 and 80% respectively, with the reductions evenly distributed across all age groups (Figures S5 and S6).

**Figure 4:**
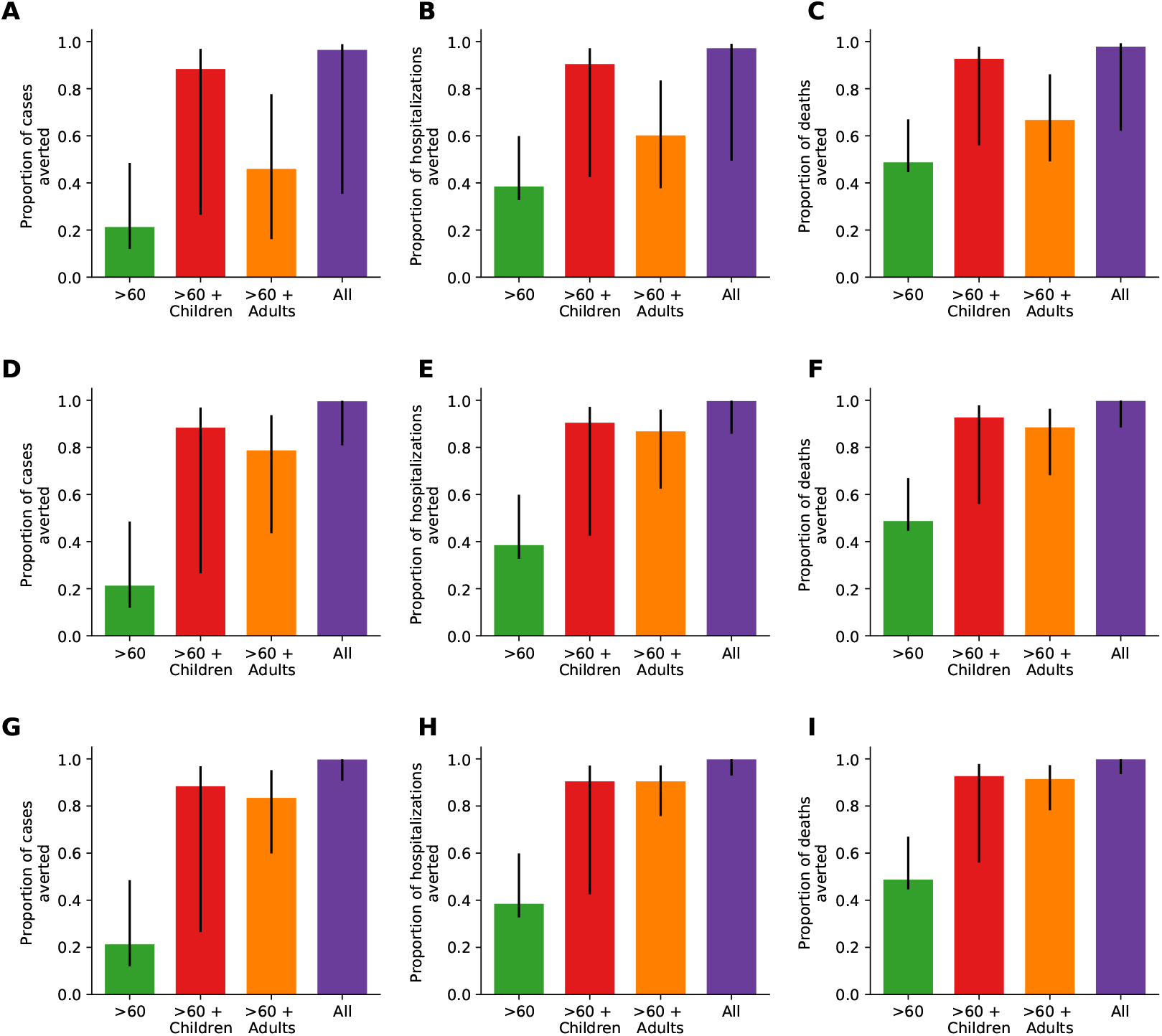
Proportion of cases (A, D, G), hospitalizations (B, E, H), and deaths (C, F, I) averted at 100 days for all scenarios considered and adults reducing their contact by (A–C) 25%, (D–F) 75%, and (G–I) 95%. (Parameter values used: *R*_0_ = 2.26, *γ* = 1*/*5.02, *σ* = 1*/*5.16)

When only 25% of adults change their contact habits, all of the interventions are set to rebound as soon as the intervention is lifted (Figure 3A). Surprisingly, cases (and hence hospitalizations and deaths) can be reduced by 94% over the first 100 days if all groups reduce their contacts with others, even when adults do so by only 25% (Figure 4A). Here, the reduction in the number of cases, hospitalizations and deaths is evenly distributed across all age groups (Figures S4A-S6A). When adults’ contacts are reduced by 75%, all of the interventions but one (where everyone reduces their contacts) rebound immediately after the end of the intervention (Figure 3B). As expected, adult groups see the greatest reductions in cases, hospitalizations and deaths from this intervention (Figures S5B and S6B). Interestingly, at this rate, the strategy that reduces the contact of adults over 60 and children averts 10% more cases than the strategy that targets adults only (87% vs. 77% cases averted) but they yield similar reductions in hospitalizations and deaths (88 and 86% of hospitalizations averted; 91 and 87% of deaths averted, respectively, Figure 4B). When adults further reduce their contacts by 95%, both strategies avert over 90% of the hospitalizations and deaths over the first 100 days (Figure 4C). Further, once adults reduce their contact rates by 75%, the strategy *all* mitigates the outbreak (Figures 3B-C, 4B-C, S4B-C-S6B-C). It is important to note that the error bars are considerably large when adults reduce their contact rates by 25%, and this uncertainty tends to smooth out as the adults further reduce their contact rates.

## 4 Discussion

The term ‘flatten the curve’, originating from the Centers for Disease Control and Prevention [28], has been used widely to describe the impact of social distancing interventions. Here, we used a mathematical model to quantify this term by measuring the short-term number of cases, hospitalization, and deaths averted over the first 100 days under four different social distancing interventions and assuming different levels of compliance in the adult population. When looking at the short-term impact (here considered to be over the first 100 days) of social distancing interventions started early on in the epidemic, our models suggest that only a social distancing intervention involving all the age groups in the population would considerably decrease the number of cases and delay the epidemic the most. However, with as little as 25% reduction in the contact rates in the adult population (combined with 95% in the older adults), one could reduce the number of hospitalizations and deaths by over 90%. Furthermore, reducing the contact rates of adults over 60 alone can have a considerable impact, reducing hospitalizations and deaths by over 30%, in agreement with what others have found [29]. It is important to interpret our results with caution. As any model, we have made important assumptions that could overestimate the effect of the interventions (see limitations below). Quantifying the short-term impact of an intervention is very important since it allows decision makers to estimate the immediate number of resources needed and to plan for further resources in the long term.

However, our simulations suggest that even in the more optimistic scenario, where all the age groups reduce their contact rates over 85%, the epidemic is set to rebound once the social distancing interventions are lifted. Our results suggest that social distancing interventions can “buy us” several weeks that could prove to be essential to strengthen our health systems and restock medical supplies, but will fail to mitigate the current pandemic. This is in agreement with other modeling results which suggested that very long periods of social distancing would be needed to control transmission [20]. Yet, sustaining social distancing interventions over a period of several months might not be feasible, both economically and socially. A combination of social distancing interventions with testing, isolation and contact tracing of new cases is needed to suppress transmission of SARS-CoV-2 [30]. Further, these interventions need to happen in synchrony around the world, as a new imported case could spark a new outbreak in any given region.

Furthermore, our results highlight the importance of the timing of social distancing interventions relative to the epidemic curve. If the intervention is put in place (and lifted) early on in the epidemic, it results in a delay of the epidemic, but if the intervention occurs later on, then it results in flattening the curve. Indeed, the effectiveness of the intervention will depend on the ratio of the number of susceptible, infected and recovered people in the population at the beginning of the intervention. There is evidence that recovered individuals will have immunity to COVID-19 but it is unclear at this point how long this immunity will last. Recovered individuals would be able to re-enter the work force and help the most vulnerable groups. It is then of paramount importance to have an accurate estimate of the number of current and recovered cases to evaluate possible interventions. As of March 12, 2020, the US had tested fewer than 15,000 CDC samples (in contrast to over 200,000 tests performed in South Korea by mid-March) [32]. It is imperative to expand the testing capabilities in all countries affected.

Our results suggest that the infectious period is a key parameter that has an extraordinary influence in the modeled speed of an epidemic and hence in the effectiveness of the interventions considered. However, current estimates of the infectious period range from 5.5 days [18] to as long as 20 days [27]. We urgently need to perform studies to have a better understanding of the duration of infectiousness of SARS-CoV-2.

Our work has several limitations and has to be interpreted accordingly. First, deterministic mathematical models tend to overestimate the final size of an epidemic. Further, deterministic models will always predict a rebound in the epidemic once the intervention is lifted provided that the number of infected people (exposed or infectious) is positive. To avoid that problem, we forced our infected compartments to be zero if they had less than one person infected at any given time. Second, we considered the latent period to be equal to the incubation period, but it has been suggested that pre-symptomatic transmission is occurring [22], and the virus SARS-CoV-2 is shed for a prolonged time after symptoms end [33]. It is unclear at this point however, if virus shed by convalescent individuals can infect others. Further, we considered that mild and severe cases would be equally infectious. In this sense, our results are conservative. We used mortality and hospitalizations data from China to assess hospitalizations and deaths rates, but these rates will presumably be country-dependent. New information about the epidemiological characteristics of SARS-CoV-2 is continuously arising, and incorporating such information in mathematical models such as ours is key to provide public health officials with the best tools to make decisions in uncertain times. Taken together, our results suggest that more aggressive approaches need to be taken to mitigate the transmission of SARS-CoV-2. Social distancing interventions need to occur in tandem with testing and contact tracing to minimize the burden of COVID-19.

## Data Availability

Full code to run the model will be available upon request.

## Acknowledgments

We thank Dr. Dobromir Dimitrov for helpful discussions. LM was supported from Fred Hutchinson Cancer Research Center research discretionary funds.

## Appendix Description of the model

We considered an age-structured SEIR model with 10 discrete age groups. The equations for the model are as follows:

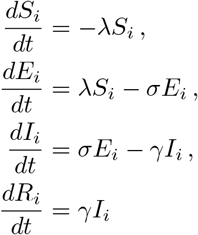

where the state variables *S*_*i*_, *E*_*i*_, *I*_*i*_, and *R*_*i*_ represent the numbers of susceptible, latent, infectious, and removed individuals in each age group *i*. Removed individuals are those who can no longer infect others. The force of infection is represented by 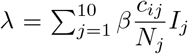, where *c*_ij_ corresponds to the estimated number of contacts per day between age class *i* and *j* in a total population size *N*. Based on current estimates [14] that put less than one percent of the infections as asymptomatic, we considered only symptomatic infections. Details of parameters values can be found in Table 1 of the main text.

**Table S1:** The delay in epidemic peak under different social distancing measures for varying infectious periods. Parentheses show the number of days delayed by each intervention compared to no intervention.

|  | 5 days<br>(Figure 2A) | 6 days<br>(Figure 2B) | 7 days<br>(Figure 2C) | 8 days<br>(Figure 2D) |
| --- | --- | --- | --- | --- |
| Do nothing | 86 | 94 | 102 | 110 |
| > 60 | 93 (+7) | 100 (+6) | 107 (+5) | 115 (+5) |
| > 60 and Children | 109 (+23) | 117 (+23) | 125 (+23) | 133 (+23) |
| > 60 and Adults | 105 (+19) | 111 (+17) | 118 (+16) | 126 (+16) |
| All | 136 (+50) | 145 (+51) | > 150 (> 48) | > 150 (> 40) |

### Additional figures and tables

**Figure S1:**
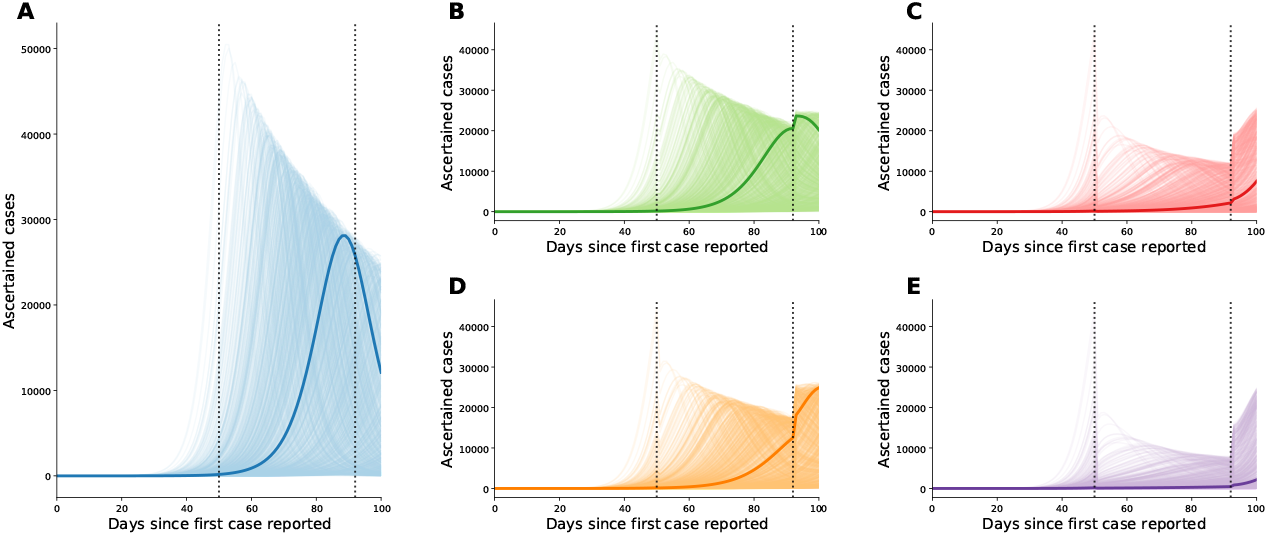
Number of ascertained cases for five scenarios of social distancing: (A) none, (B) *>* 60 only, (C) *>* 60 and children, (D) *>* 60 and adults, and (E) all. Thin lines show 1000 realizations of varying parameters for *R*_0_, *σ* and *γ* (see Methods). Thick lines show the median simulation, corresponding to *R*_0_ = 2.26, *γ* = 1*/*5.015 and *σ* = 1*/*5.159. Here, we assumed that adults over 60 will reduce their contact by 95%, children by 85% and adults by 25%. The dotted lines represent the beginning and end of the social distancing intervention, at 50 and 92 days respectively.

**Figure S2:**
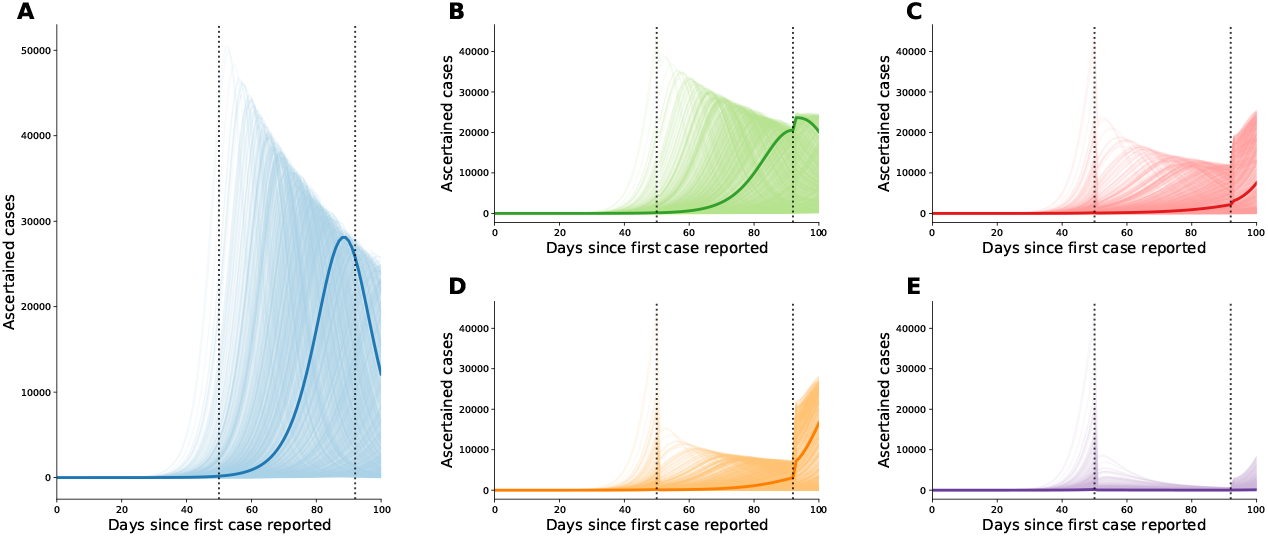
Number of ascertained cases for five scenarios of social distancing: (A) none, (B) *>* 60 only, (C) *>* 60 and children, (D) *>* 60 and adults, and (E) all. Thin lines show 1000 realizations of varying parameters for *R*_0_, *σ* and *γ* (see Methods). Thick lines show the median simulation, corresponding to *R*_0_ = 2.26, *γ* = 1*/*5.015 and *σ* = 1*/*5.159. Here, we assumed that adults over 60 will reduce their contact by 95%, children by 85% and adults by 75%. The dotted lines represent the beginning and end of the social distancing intervention, at 50 and 92 days respectively.

**Figure S3:**
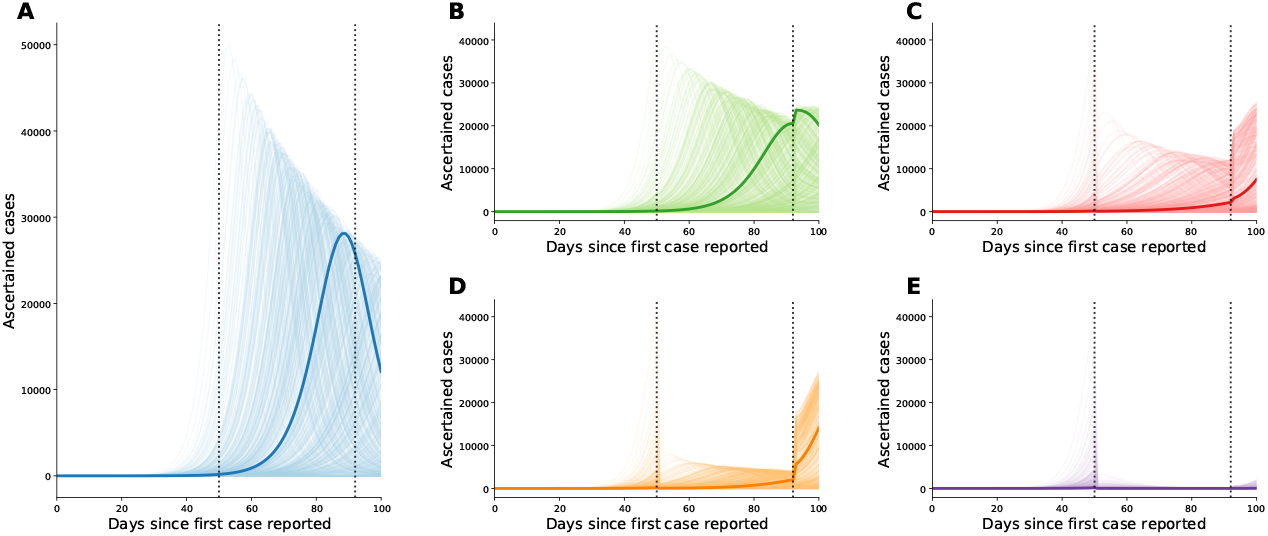
Number of ascertained cases for five scenarios of social distancing: (A) none, (B) *>* 60 only, (C) *>* 60 and children, (D) *>* 60 and adults, and (E) all. Thin lines show 1000 realizations of varying parameters for *R*_0_, *σ* and *γ* (see Methods). Thick lines show the median simulation, corresponding to *R*_0_ = 2.26, *γ* = 1*/*5.015 and *σ* = 1*/*5.159. Here, we assumed that adults over 60 will reduce their contacts by 95%, children by 85% and adults by 95%. The dotted lines represent the beginning and end of the social distancing intervention, at 50 and 92 days respectively.

**Figure S4:**
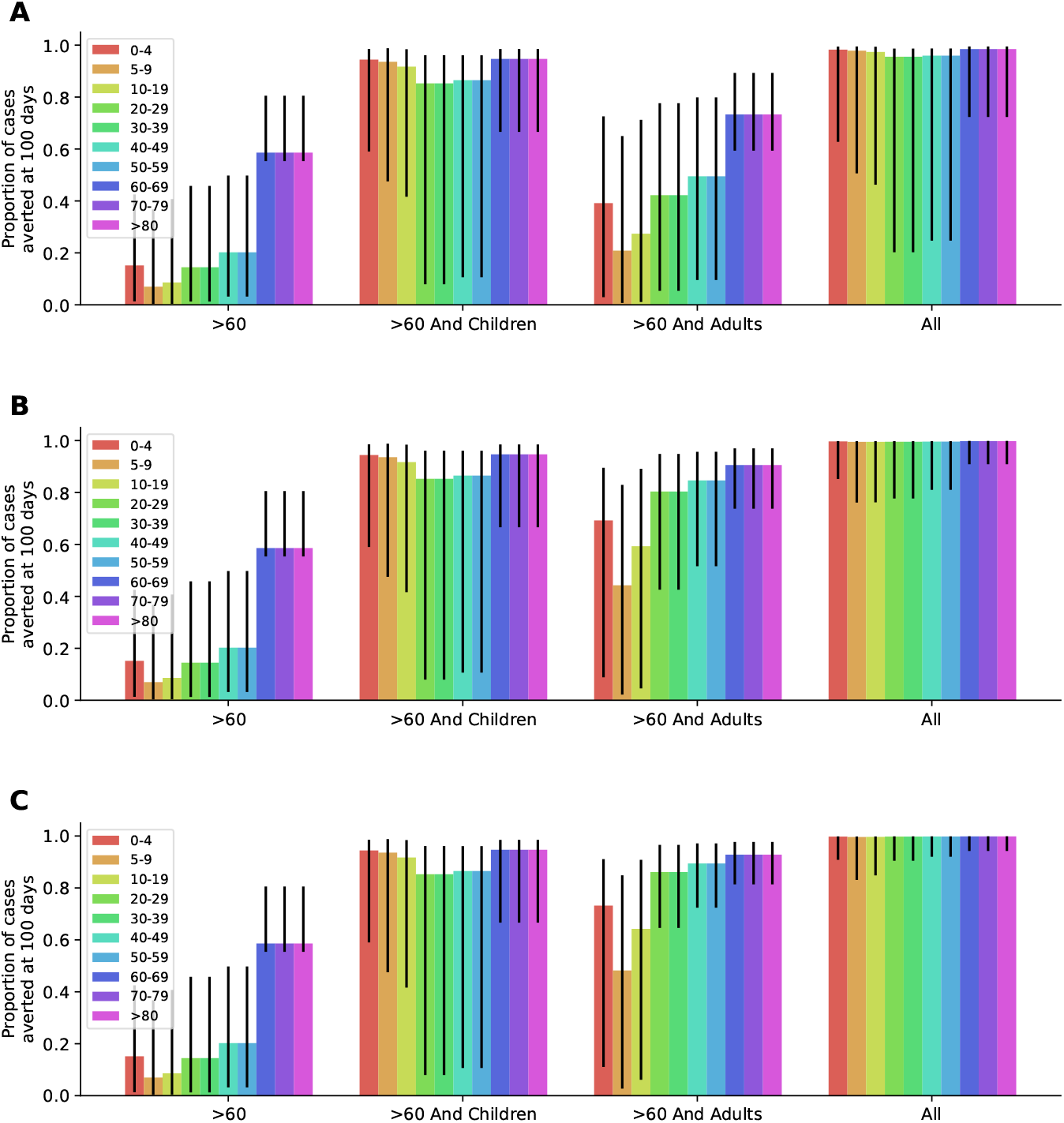
Proportion of cases averted at 100 days under four scenarios of adults reducing their contact by (A) 25%, (B) 75%, and (C) 95%.

**Figure S5:**
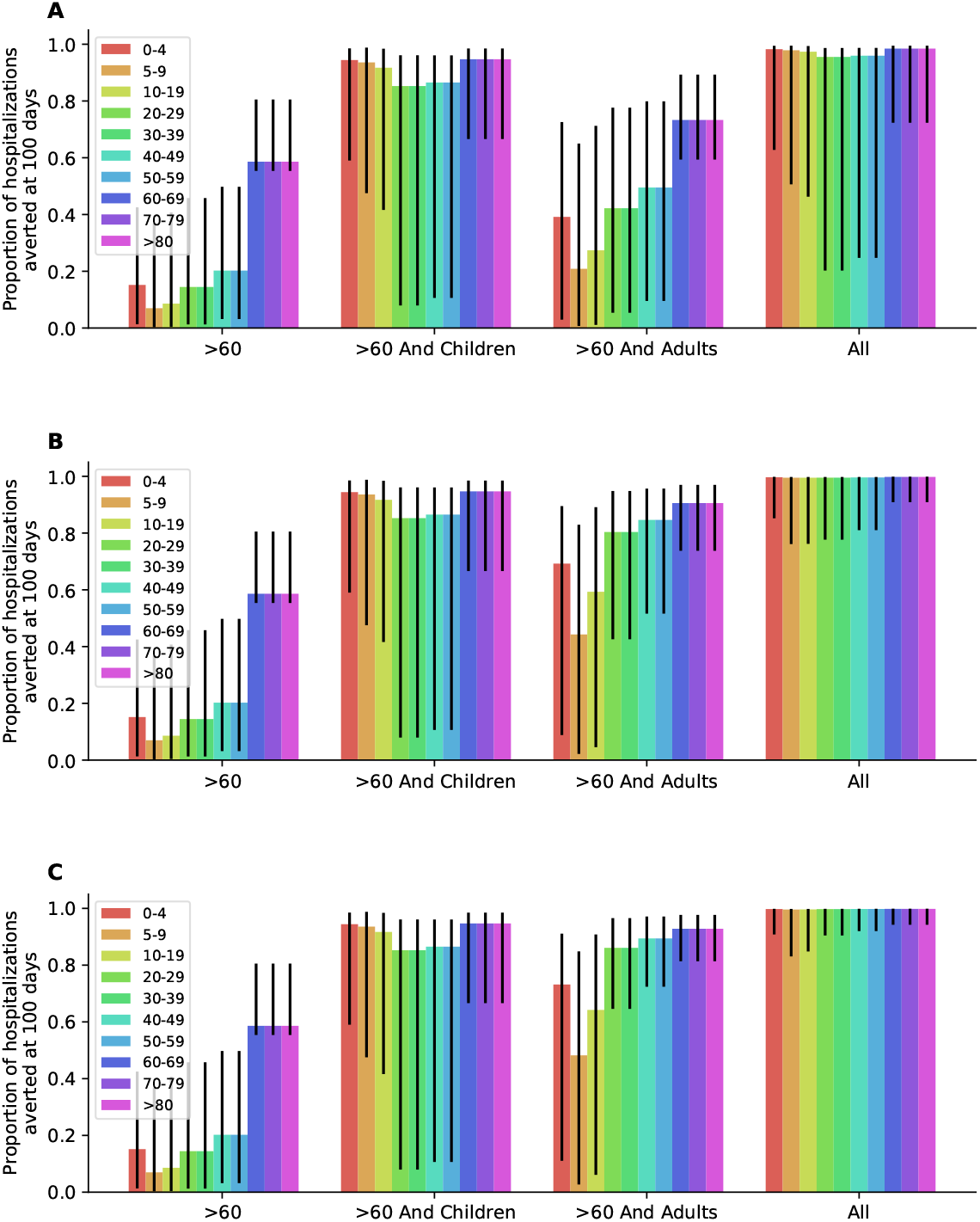
Proportion of hospitalizations averted at 100 days under four scenarios of adults reducing their contact by (A) 25%, (B) 75%, and (C) 95%.

**Figure S6:**
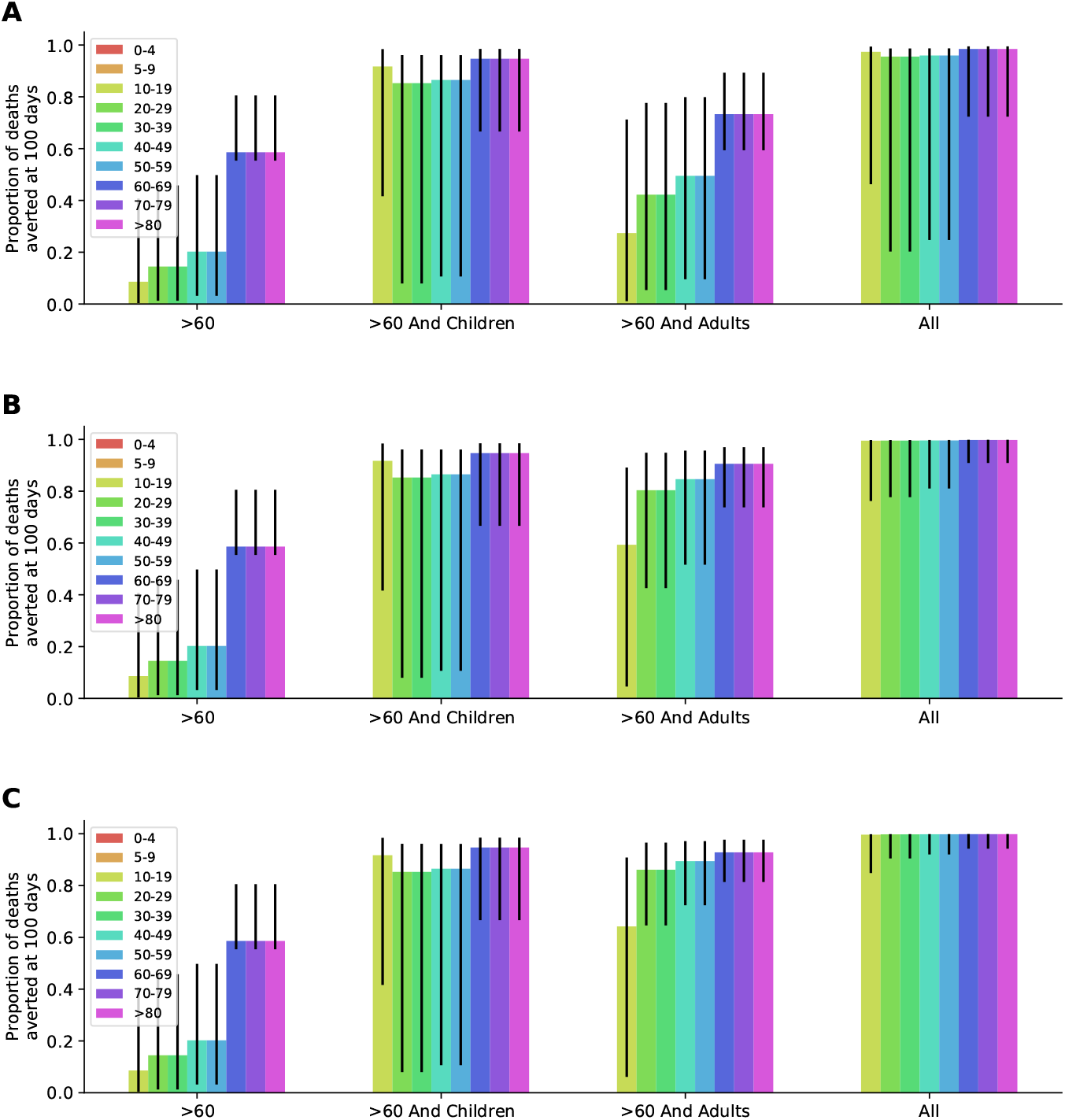
Proportion of deaths averted at 100 days under four scenarios of adults reducing their contact by (A) 25%, (B) 75%, and (C) 95%.

## Notes

### Competing Interest Statement

The authors have declared no competing interest.

### Funding Statement

This work was supported from Fred Hutchinson research discretionary funds.

